# Association between viral infections and Ménière’s disease risk: a two-sample bidirectional Mendelian randomization study and meta-analysis

**DOI:** 10.1101/2025.09.09.25335399

**Authors:** Xiaofei Li, Yifan Yu, Xiaoyang Li, Fan Jiang, TongTong Zhang, Jiamin Lu, Na Li, Haibo Wang, Hongkai Li, Daogong Zhang

## Abstract

**Introduction:** Meniere’s disease (MD) is a chronic inner ear disorder that can lead to hearing and balance disabilities, yet its etiology remains unknown. We employed Mendelian randomization (MR) analysis to investigate the causal relationship between viral infections and MD.

**Methods:** We employed bidirectional two-sample MR analysis to investigate the causal relationship between 13 viral infections and MD. Single nucleotide polymorphisms (SNPs) were selected as instrumental variables for MR analysis from large European population-based genome-wide association studies (GWAS) corresponding to the 13 viral infections and Meniere’s disease. The inverse variance-weighted (IVW) method was used as the primary MR analysis approach, with sensitivity analyses conducted to assess pleiotropy and heterogeneity. Additionally, for viral infections with multiple GWAS data sources, MR analysis was performed separately for each dataset, and the results were combined using a random-effects model for meta-analysis.

**Results:** After Bonferroni correction and sensitivity analyses, MR and meta-analysis results indicated a negative causal association between genetically predicted HSV infection and Meniere’s disease (OR=0.90, CI=0.85, 0.96), as well as a negative causal effect of genetically predicted HPV virus on Meniere’s disease (OR=0.82, CI=0.75, 0.90). There was no evidence of a causal relationship from Meniere’s disease to viral infections.

**Conclusions:** Our research findings offer compelling evidence that genetically predicted HPV and HSV infections are associated with a reduced risk of MD. These discoveries contribute to an enhanced understanding of the etiology of MD and the development of future interventions to mitigate the risk of MD.

## Introduction

Ménière’s disease (MD) is a clinical condition characterized by spontaneous vertigo attacks, along with sensorineural hearing loss, tinnitus and ear fullness in the affected ear. The pathogenesis of this disease remains unclear(Nakashima et al., 2016). MD affects approximately 50 to 200 per 100,000 adults, with the most common age group being between 40 to 60 years old(Basura et al., 2020). MD can cause significant functional disability for patients, as they deal with ongoing balance and auditory issues(Cohen et al., 1995). This also leads to a heavy emotional burden for those affected. The average annual cost per person for managing MD is estimated to be between £3341 (US $4421.65) and £3757 (US $4972.21), which is higher than the costs associated with managing asthma and migraine(Tyrrell et al., 2016). Therefore, identifying the cause of MD is crucial as it can potentially lead to preventative measures to reduce the incidence of the disease. It may also allow for targeted screening of individuals who are at a higher risk of developing MD. While previous studies have attempted to find the cause of MD, a clear etiology has yet to be identified.

There are multiple theories aimed at explaining the pathophysiology and origins of MD(Nakashima et al., 2016; Rizk et al., 2022). One theory proposes a potential viral component in the development of the disease(Kurtzman and Sioshansi, 2023), with viruses such as herpes simplex virus (HSV), cytomegalovirus (CMV), Epstein Barr virus (EBV), influenza virus, adenovirus (AV), Coxsackie virus B, measles, and Varicella Zoster virus (VZV) being implicated. However, not all studies support this viral link. A recent meta-analysis of 14 case-controlled observational studies, involving 611 MD patients and 373 controls, examined the association between viral infections and MD(Dean et al., 2019). The analysis found a small correlation between CMV infection and MD, but no evidence of a connection between MD and HSV-1, HSV-2, VZV, or EBV. Furthermore, attempts to use antivirals as a treatment for Meniere’s disease have shown controversial results(Cao et al., 2019; Syed et al., 2015). These observational studies are limited by small sample sizes and significant heterogeneity in selection, matching, and ascertainment methods related to viral infection. Therefore, future research efforts should focus on generating more robust and high-quality evidence to validate these findings. A comprehensive study with a rigorous approach is urgently needed to establish a definitive conclusion regarding the causal relationship between viral infection and MD.

In this study, Mendelian Randomization (MR) aims to utilize genetic variants as instrumental variables (IVs) to infer the causal relationship between 13 viruses exposure and outcome of MD. Crucially, these genetic variants employed as IVs are not required to be direct pathogenic mutations of the MD but a genetic susceptibility association of 13 viruses that have either been studied previously through traditional observational research or have not yet been explored on MD, including VZV, HSV, CMV, human papilloma virus(HPV), EBV, human papilloma virus(HIV), hepatitis B virus (HBV), rubella virus(RV), Measles virus (MV), and infuenza virus (IV), Mumps, COVID-19 and poliovirus. The main aim of this MR analysis was to elucidate the direction and strength of the causal relationship between viral infection and the risk of developing MD.

## Methods

### Design

This study employed a bidirectional two-sample Mendelian randomization (MR) design to investigate the causal relationships between 13 viral infections, including CMV, COVID-19, EBV, HBV, HIV, HPV, HSV, Influenza, Mumps, Measles, Poliovirus, Rubella virus, and VZV, and MD. For the MR analysis to be valid, the study adhered to the following three core assumptions(Emdin et al., 2017): (1) genetic instruments are strongly associated with the exposure, (2) genetic instruments are independent of confounders, and (3) genetic instruments affect the outcome solely through the exposure. This study followed the Strengthening the Reporting of Observational Studies in Epidemiology Using Mendelian Randomization (STROBE-MR) guidelines (Figure 1)(Skrivankova et al., 2021).

**Figure 1.**
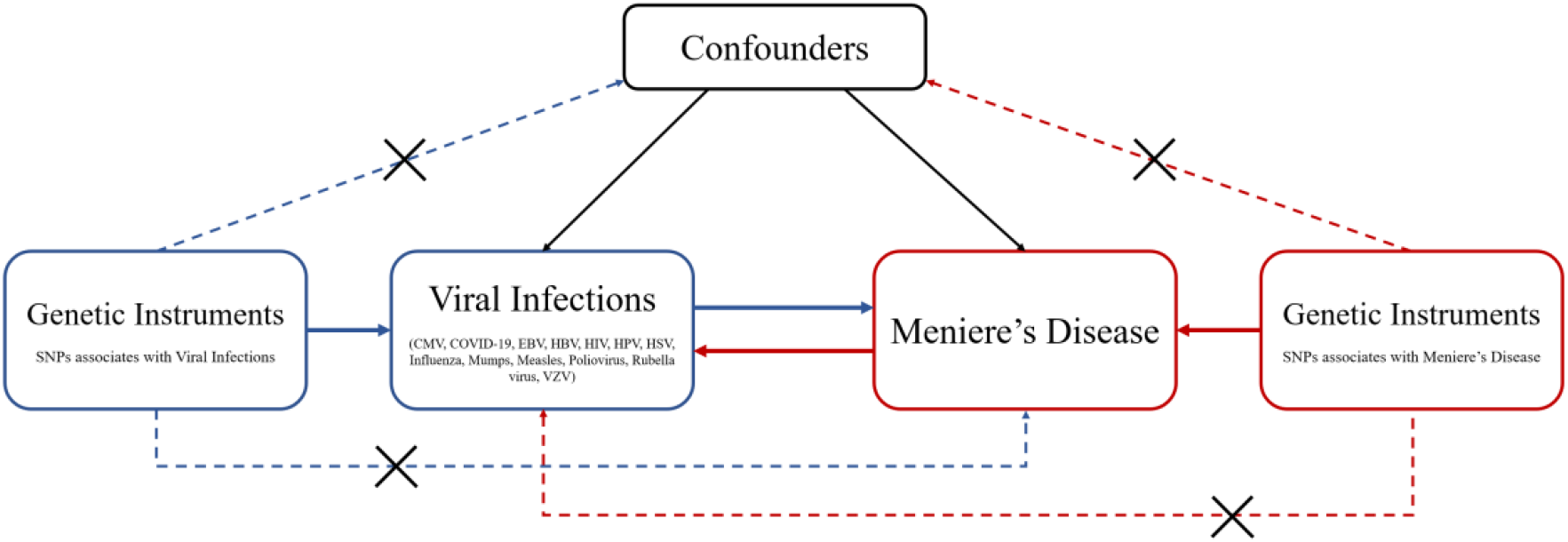
Basic assumptions of Mendelian randomization.

### Data Sources

This study incorporated 17 GWAS datasets covering 13 viral infections, along with one GWAS dataset for Meniere’s disease. For FinnGen datasets(Kurki et al., 2023), the diagnosis of viral infections is established through the utilization of International Classification of Diseases (ICD) codes within the Finnish Register of Inpatients, Outpatients and Causes of Death. For **Varicella Zoster Virus (VZV)**, three separate GWAS datasets were included: UK Biobank (GCST90018941) with 522 cases and 351,740 controls(Sakaue et al., 2021), FinnGen dataset for Zoster, which comprised 2,080 cases and 211,856 controls, and FinnGen dataset for Varicella, including 710 cases and 211,856 controls. For **Herpes Simplex Virus (HSV)**, two GWAS datasets were sourced from the FinnGen database: Anogenital herpesviral infection dataset included 891 cases and 212,952 controls, and Herpesviral infections dataset included 1,595 cases and 211,856 controls. The GWAS dataset for **Cytomegalovirus (CMV)** was obtained from FinnGen, with 270 cases and 213,666 controls. For **Human Papillomavirus (HPV)**, the GWAS dataset was derived from Suhre et al(Suhre et al., 2017). (2017), consisting of 997 participants. The GWAS dataset for hospitalized **COVID-19** was sourced from the COVID-19 Host Genetics Initiative(2020), comprising 9,986 cases and 1,877,672 controls. For **Mumps**, the dataset was obtained from FinnGen, including 680 cases and 333,715 controls. For **Epstein-Barr Virus (EBV)**, the GWAS dataset from FinnGen comprised 2,099 cases and 333,715 controls. For **Poliovirus**, the GWAS dataset from FinnGen included 230 cases and 217,637 controls. The GWAS dataset for **Human Immunodeficiency Virus (HIV)** was sourced from FinnGen, with 357 cases and 218,435 controls. For **Hepatitis Virus**, data were obtained from two sources: UK Biobank(Sakaue et al., 2021), with 145 cases and 351,740 controls, and FinnGen dataset, which included 1,226 cases and 217,566 controls. For **Rubella Virus (RuV)**, the dataset from FinnGen included 553 cases and 211,856 controls. For **Measles**, the FinnGen GWAS dataset comprised 180 cases and 211,856 controls. For **Influenza**, the dataset from FinnGen included 4,262 cases and 188,868 controls. Finally, the GWAS dataset for **Meniere’s Disease** was obtained from FinnGen, with 1,151 cases and 209,582 controls. All GWAS datasets were derived from European populations (Appendix Table S1).

### Genetic Instruments

To ensure robust genetic instruments across all analyses, we selected variants from the GWAS datasets using genome-wide suggestive significance thresholds (*p* < 5 × 10^−6^). We applied an r^2^ threshold of 0.001 (1000 kb) to account for linkage disequilibrium (LD) between SNPs, minimizing bias in the MR estimates. Additionally, we calculated the F-statistic to avoid weak instrument bias. The F-statistic was calculated as: F = *R*^2^ × (*N* − *K* − 1)/*K* × (1 − *R*^2^), where R^2^ represents the proportion of exposure variance explained by the SNPs, K is the number of instruments, and N is the sample size from the corresponding GWAS. An F-statistic greater than 10 indicates that the SNPs possess strong statistical power, reducing the likelihood that the MR analysis results will be biased due to weak instruments(Burgess et al., 2017). This suggests that the selected SNPs can be considered valid instrumental variables for analysis. The SNPs used in this study are shown in Appendix Table S2.

### Statistical Analysis

We performed MR analysis using the *TwoSampleMR* package in R to investigate causal relationships between viral infections and MD(Hemani et al., 2017). The inverse-variance weighted (IVW) method was the primary MR approach to estimate causal effects(Burgess et al., 2017), with Bonferroni correction applied to account for multiple comparisons (adjusted significance level *p* < 0.05/17 = 0.003, based on 17 viral infection datasets). We also conducted sensitivity analyses using methods including simple median, weighted median(Bowden et al., 2016), MR-Egger(Bowden et al., 2015), MR-robust adjusted profile score (MR-RAPS), and Mendelian Randomization Pleiotropy Residual Sum and Outlier (MR-PRESSO) to verify the robustness of the findings(Verbanck et al., 2018). The MR-Egger intercept was used to detect horizontal pleiotropy, and heterogeneity was assessed using the Cochran’s Q test, with random-effects IVW models applied where necessary. Leave-one-out analysis was employed to ascertain whether the results were influenced by a single SNP. To explore potential bidirectional relationships, we performed two MR analyses: first, treating viral infections as the exposure and MD as the outcome, and second, reversing the roles. For viral infections such as HSV and VZV, with multiple GWAS datasets, we conducted a meta-analysis of the IVW estimates using the random-effects model. All statistical analyses were performed using R version 4.3.2. The MR analyses were conducted using the R packages *TwosampleMR, mr*.*raps*, and *MRPRESSO*, while the meta-analyses were carried out using the *meta* package(Balduzzi et al., 2019).

## Results

### Causal Association of Viral Infections with MD

The primary MR results investigating the causal effects of viral infections on MD are presented in Figure 2. The IVW analysis for HSV, based on the anogenital herpes viral infection GWAS, showed a negative causal association with MD (OR=0.88; 95% CI=0.80, 0.96), supported by eight SNPs. A second HSV GWAS (herpes viral infections) suggested a potential negative causal association (OR=0.93, p=0.083). Meta-analysis of the two HSV datasets further confirmed a significant negative causal effect (OR=0.90, CI=0.85, 0.96) at the Bonferroni-adjusted significance level (p < 0.003), indicating a robust association between HSV infection and a lower risk of MD. Additionally, HPV changed (1 standard deviation) was found to have a negative causal effect on MD (OR=0.82, CI=0.75, 0.90), also meeting the Bonferroni-adjusted threshold (p < 0.003). Hepatitis virus showed a suggestive causal relationship (OR=0.97, p=0.044), but it did not reach the Bonferroni significance threshold, as well as the suggestive causal relationship between HIV and MD (OR=0.97, p=0.039). Other viral infections, including CMV, COVID-19, EBV, Influenza, Mumps, Measles, Poliovirus, Rubella virus, and VZV, showed no significant causal associations with MD (p > 0.05).

**Figure 2.**
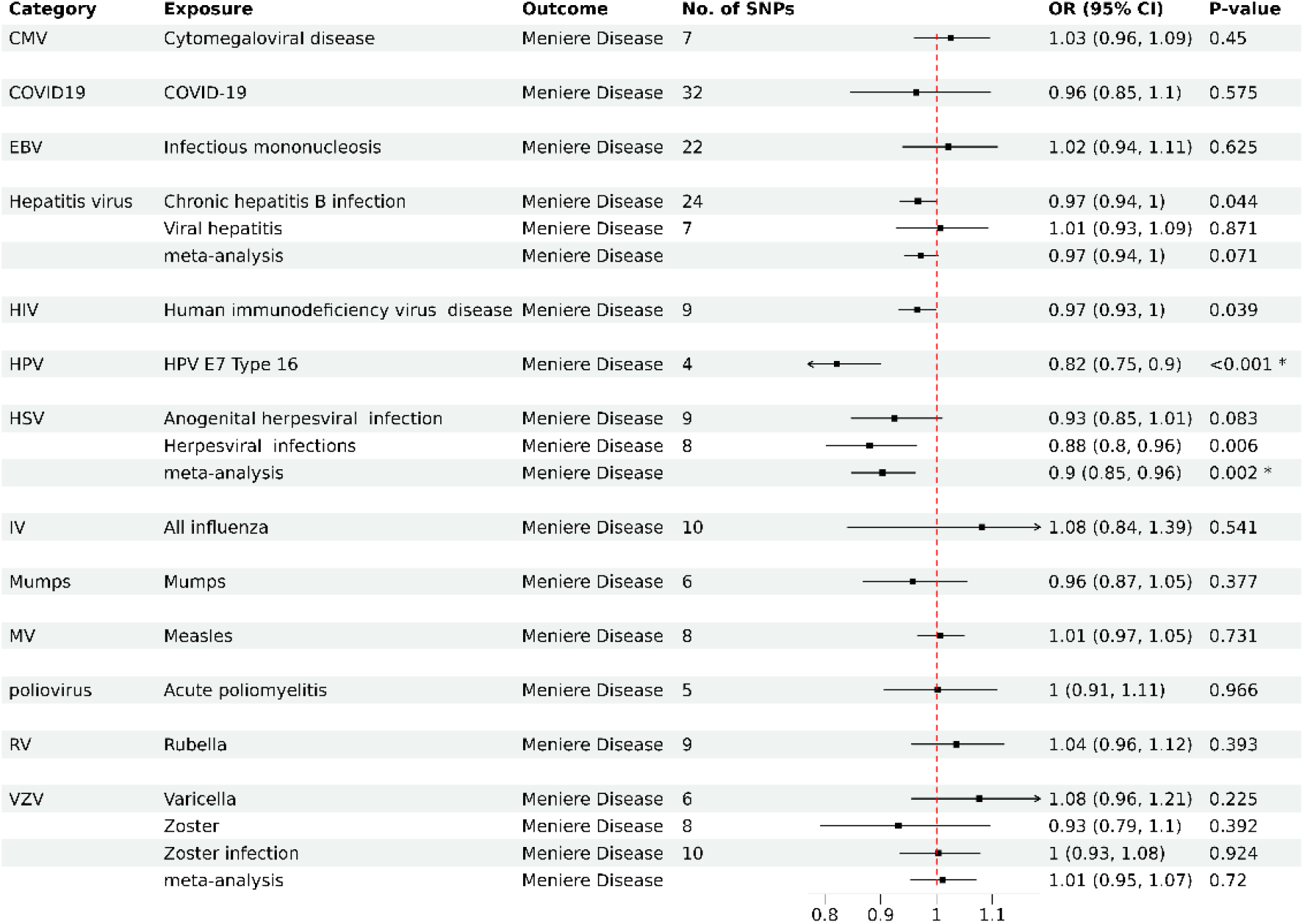
Odds ratios for associations between genetically predicted viral infection and MD. ^*^Bonferroni-adjusted significance level.

The results were consistent across sensitivity analyses, with the HPV-MD association supported by both simple median and MR-RAPS methods, confirming the robustness of the findings. The MR-Egger intercept test indicated no evidence of directional pleiotropy, and heterogeneity tests along with MR-PRESSO analyses showed no significant heterogeneity for HPV or HSV. The F-statistics for all exposed instrumental variables were greater than 10 (ranging from 21.73 to 39.78), indicating a low likelihood of bias in the results due to weak instruments and sample overlap. The results of all MR methods, as well as the results of sensitivity analyses, are presented in Appendix Table S3 and Figure S1-3.

### Causal Association of Meniere’s Disease with Viral Infections

MR analysis of the reverse direction, evaluating the causal effect of MD on viral infections, showed no significant associations for any of the 13 viral infections based on the IVW method (Figure 3). Other MR methods corroborated these null findings. The MR-Egger intercept test revealed no evidence of pleiotropy, and heterogeneity tests indicated no significant heterogeneity across SNPs. Results from MR-PRESSO, after excluding potential outlier SNPs, remained consistent. The F-statistic for MD instruments was 24.09, suggesting a low likelihood of weak instrument bias. Overall, these results provide robust evidence for the absence of a causal relationship between MD and viral infections. The results of all MR methods, as well as the results of sensitivity analyses, are presented in Appendix Table S4 and Figure S4-6.

**Figure 3.**
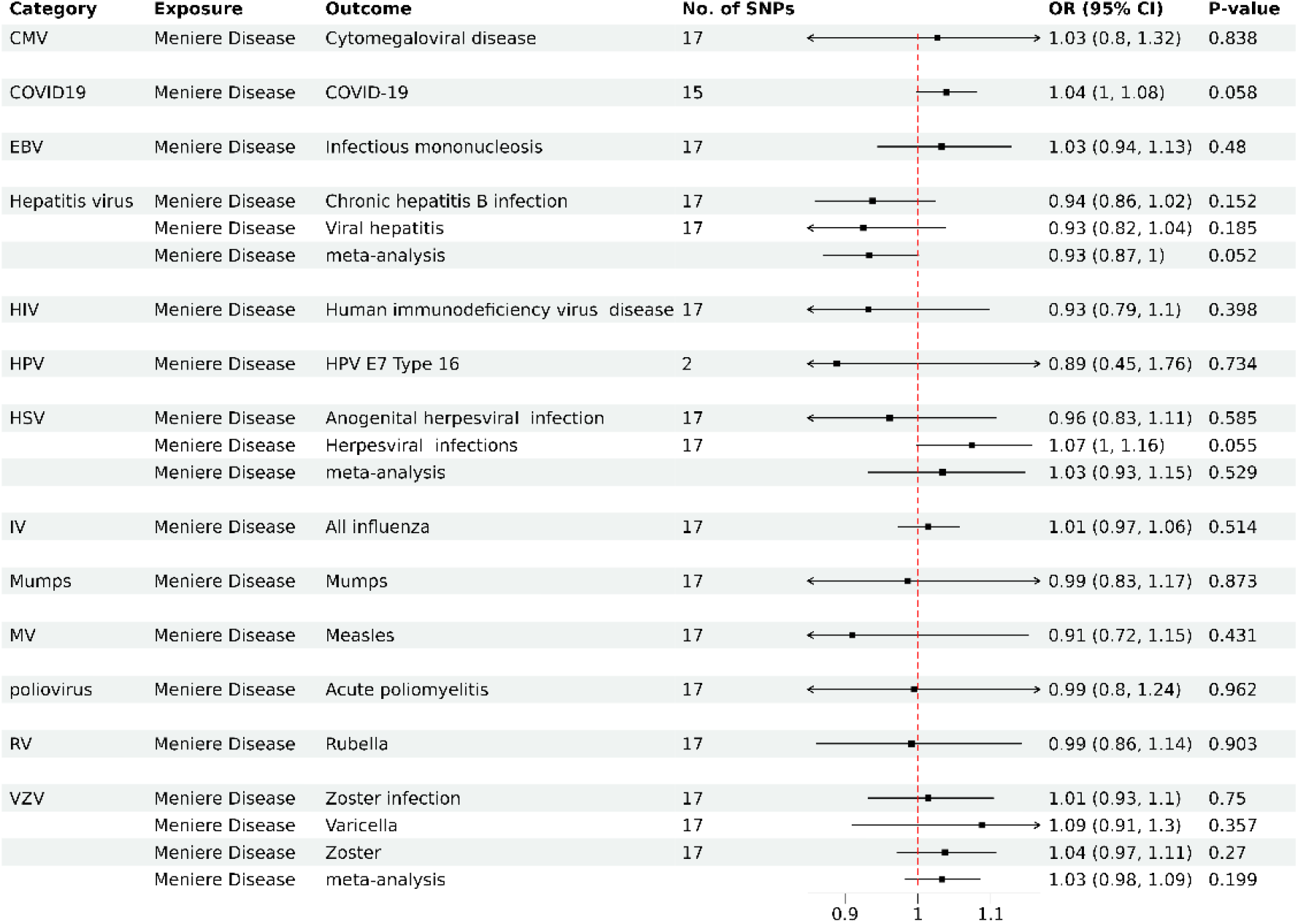
Odds ratios for associations between genetically predicted MD and viral infection.

## Discussion

Viral infections have been suggested as potential risk factors for MD, but the findings have been controversial. As far as we know, this is the most comprehensive analysis examining the causal relationships between 13 distinct virus infections and MD. Our results reveal a notable association between herpes simplex virus (HSV) and MD, along with an association between human papillomavirus (HPV) and MD. The result is less susceptible to confounding and reverse causality bias than many previous conventional observational studies(Davies et al., 2018).

Numerous studies have investigated the potential viral causes of MD, such as HSV, CMV, EBV, VZV, AV, Coxsackie virus B, measles virus (MV), and influenza virus. Various specimens, including serum, PBMC, endolymphatic sac, perilymph fluid, and vestibular ganglion, have been tested using different detection methods such as viral titers, antibody reactivity, TEM, and viral DNA analysis in labyrinthine or retro-labyrinthine tissue of MD patients. However, results have been inconclusive and not consistently reproducible, leading to ongoing debates about the viral etiology of MD. A previous study reported HSV is more commonly isolated from vestibular ganglia of patients with MD than the general population by nested polymerase chain reaction while another study reported no viral DNA was detected in the vestibular ganglion of all 7 patients with definite MD included(Vrabec, 2003). D B Welling et al reported only 2 of the 22 extracts from the endolymphatic sacs was detected HSV DNA an no other virus evidence obtained from patients with MD(Welling et al., 1994). On the contrary, I K Arenberg et al reported that a very high portion (77.8%) of MD patients were detect CMV in endolymphatic sac tissues with a significant difference compare to the controls(Arenberg et al., 1997). In larger studies involving blood specimens, elevated antibody titers against certain viruses have been observed in MD patients compared to controls, suggesting a potential viral link to MD(Pyykko and Zou, 2008). However, a recent meta-analysis found only a small association between CMV infection and MD(Dean et al., 2019), with no associations found for other viruses like HSV-1, HSV-2, VZV, or EBV. Therefore, the current evidence is insufficient to definitively establish a clear and comprehensive relationship between viral infections and MD, as the presence of viral DNA or anti-virus antibody does not necessarily indicate a causal association to the disease process.

Mendelian randomization (MR) is a powerful method that uses genetic variation as an instrumental variable to assess the association between exposure and disease(Bowden and Holmes, 2019). MR analysis helps to reduce confounding and reverse causality, making it increasingly used to establish a causal relationship between potentially modifiable risk factors and outcomes. However, the causal relationship between viral infections and MD remains poorly understood and unestablished. In this study, we aimed to address the following questions: 1) Is viral infection associated with MD risk? 2) Which types of viruses are involved in this association? 3) What is the directionality of this association? Our findings provide valuable insights into these questions. Interestingly, our results suggest that HSV and HPV infections may actually reduce the risk of MD.

This unexpected finding is not unprecedented, as protective effects of certain viral infections have been reported in other diseases. For example, studies have shown that infection with CMV and EBV can induce apoptosis of immune cells, leading to a weakened immune response and slowing the progression of Celiac Disease(Lerner et al., 2017). Additionally, VZV infection has been found to significantly reduce the risk of developing low-grade gliomas by recruiting NK cells and T lymphocytes(Gerada et al., 2020; Nikzad et al., 2019), increasing inflammatory factors, and altering the overall immune response. These findings suggest that viral infections may have a protective effect on the host in certain circumstances. This protective crosstalk between viruses and the immune system may reflect a mutually beneficial balance between two symbiotic ecosystems. This novel perspective offers new insights into the complex interactions between viral infections and the immune system of MD patients.

HPV is a double-stranded circular DNA virus that belongs to the papillomavirus family. Transmission usually occurs through skin-to-skin or mucosa-to-mucosa contact, with the virus entering the body through cutaneous or mucosal trauma. Globally, approximately 50% of both men and women are likely to experience HPV infection at least once in their lifetime(Handler et al., 2015). While extensive research has focused on the role of HPV in cancer development, previous studies have not reported any association between HPV and MD. On the other hand, HSV-1 and HSV-2 are highly prevalent human pathogens with global prevalence levels of approximately 67% and 13%(James et al., 2020), respectively. Primary HSV infection typically occurs in childhood, with the virus later remaining latent in nerve ganglia. Upon reactivation, the virus travels along nerve fibers to the skin or mucous membranes, resulting in a new infection(Gopinath et al., 2023). Despite numerous studies suggesting an association between HSV and MD, a causal relationship has not been established. Furthermore, these studies often indicate that HSV is a risk factor for MD.

Our study has uncovered a novel finding that HPV and HSV are associated with a reduced risk of MD, supported by robust results following p-value correction and sensitivity analysis. However, the exact biological mechanisms through which HPV and HSV reduce the risk of MD remain unclear. We suggest three hypotheses to explain this phenomenon. Firstly, virus infections may modulate the immune response. HPV can regulate NK cell activity, regulate dendritic cell activation of T cells, and thereby modulate the host’s immune response(Bashaw et al., 2017; Zhou et al., 2019). Similarly, HSV can suppress the response of monocytes to interferon-gamma, dampens the inflammatory response, and changes the cytokine profile and chemokine response(Albers et al., 1989). Additionally, HSV influences the activation and proliferation of T cell subsets and promotes antigen presentation in dendritic cells(Gurczynski and Moore, 2020; Yu et al., 2018). These immune cell populations are ever reported to be significantly altered in MD patients(Flook et al., 2023; Zhang et al., 2023). Secondly, HPV and HSV may suppress inflammatory responses by influencing cytokine release. This suppression could be a strategy employed by virus to evade the host immune system(Britto et al., 2020), making it harder for the host to recognize and clear infections, yet unexpectedly reducing the risk of MD. HSV has been reported to inhibit the activation of tumor necrosis factor α and NF-kB(Zhang et al., 2013), both of which play key roles in inner ear inflammation and disease progression in MD(Huang et al., 2022; Kouhi et al., 2021; Zhang et al., 2023). Lastly, HPV and HSV infections might stimulate the production of substances such as nerve growth factors, aiding in nerve cell growth and repair. The interaction between HSV-2’s glycoprotein G and nerve growth factors could promote nerve growth(Kropp et al., 2020). The extracellular vesicles from HPV-positive cervical cancer cell lines can stimulate nerve growth(Anand et al., 1995; Lucido et al., 2019). This indicates that HPV and HSV may potentially stimulate the growth and repair of neurons through specific mechanisms, with nerve damage being identified as a pathological alteration in MD(Spencer et al., 2002; Wang et al., 2019). In total, these unexpected findings shed new light on the potential protective effects of certain viral infections and provide a novel perspective on the immune response in MD patients.

Our study has several strengths. First, this is the first study to draw causal conclusions and eliminate confounding factors and reverse causality using the two-sample MR method to investigate the relationship between viral infection and MD. Second, we included novel viruses never studied before, such as poliovirus and HPV infection. However, some limitations in this research need to be noted. Firstly, to minimize population differences, all data used in this study were derived from European population, which may limit the generalizability of the findings to other populations. Secondly, there is potential bias due to sample overlap between the viral infection GWAS data and the MD GWAS data. However, the F-statistics for the instrumental variables selected in this study were greater than 10, indicating that the SNPs used as instruments are robust(Burgess et al., 2016). Based on previous research, the likelihood of bias due to sample overlap is relatively low. Thirdly, while the results suggest a negative causal association between viral infections and MD, the underlying mechanisms by which viral infections may influence MD remain unclear and require further experimental validation.

## Conclusions

Our research findings offer compelling evidence that genetically predicted HPV and HSV infections are associated with a reduced risk of MD. These discoveries contribute to an enhanced understanding of the etiology of MD and the development of future interventions to mitigate the risk of MD.

## Data Availability

All data produced in the present study are available upon reasonable request to the authors

## Declarations

### Ethics approval and consent to participate

Not applicable.

### Consent for publication

All authors have agreed on the consent of the manuscript.

### Availability of data and materials

No datasets were generated or analysed during the current study.

### Authors’ contributions

Haibo Wang attests that all listed authors meet authorship criteria and that no others meeting the criteria have been omitted. All the authors were responsible for the decision to submit the manuscript. Concept and design: Daogong Zhang, Hongkai Li, Haibo Wang, Xiaofei Li, Yifan Yu. Acquisition of data: Yifan Yu, Xiaofei LI; Formal analysis and Interpretation of data: Xiaofei Li, Yifan Yu Hongkai Li, Daogong Zhang, Haibo Wang. Statistical analysis: Yifan Yu, Xiaofei Li, Xiaoyang Li; Drafting of the manuscript: Xiaofei Li, Yifan Yu, Hongkai Li; Critical revision of the manuscript for important intellectual content: Daogong Zhang, Hongkai Li, Haibo Wang, Fan Jiang, Na Li.

## Acknowledgments

**None**

